# De-novo FAIRification via an Electronic Data Capture system by automated transformation of filled electronic Case Report Forms into machine-readable data

**DOI:** 10.1101/2021.03.04.21250752

**Authors:** Martijn G. Kersloot, Annika Jacobsen, Karlijn H.J. Groenen, Bruna dos Santos Vieira, Rajaram Kaliyaperumal, Ameen Abu-Hanna, Ronald Cornet, Peter A.C. ‘t Hoen, Marco Roos, Leo Schultze Kool, Derk L. Arts

**Affiliations:** Amsterdam UMC, University of Amsterdam, Department of Medical Informatics, Amsterdam Public Health Research Institute, Amsterdam, The Netherlands; Castor EDC, Amsterdam, The Netherlands; Department of Human Genetics, Leiden University Medical Center, Leiden, The Netherlands; Department of Medical Imaging, Radboud Institute for Health Sciences, Radboud university medical center, Nijmegen, The Netherlands; Center for Molecular and Biomolecular Informatics, Radboud Institute for Molecular Life Sciences, Radboud university medical center, Nijmegen, The Netherlands

**Author notes:** Corresponding author at Amsterdam UMC, University of Amsterdam, Department of Medical Informatics, Amsterdam Public Health Research Institute Room J1B-109, PO Box 22700, 1100 DE, Amsterdam, The Netherlands Email address (Martijn G. Kersloot).

**Keywords:** electronic Case Report Forms, FAIR Data, machine-readable data, interoperability, patient registry

## Abstract

**Introduction:** Existing methods to make data Findable, Accessible, Interoperable, and Reusable (FAIR) are usually carried out in a post-hoc manner: after the research project is conducted and data are collected. De-novo FAIRification, on the other hand, incorporates the FAIRification steps in the process of a research project. In medical research, data is often collected and stored via electronic Case Report Forms (eCRFs) in Electronic Data Capture (EDC) systems. By implementing a de-novo FAIRification process in such a system, the reusability and, thus, scalability of FAIRification across research projects can be greatly improved. In this study, we developed and implemented a novel method for de-novo FAIRification via an EDC system. We evaluated our method by applying it to the Registry of Vascular Anomalies (VASCA).

**Methods:** Our EDC and research project independent method ensures that eCRF data entered into an EDC system can be transformed into machine-readable, FAIR data using a semantic data model (a canonical representation of the data, based on ontology concepts and semantic web standards) and mappings from the model to questions on the eCRF. The FAIRified data are stored in a triple store and can, together with associated metadata, be accessed and queried through a FAIR Data Point. The method was implemented in Castor EDC, an EDC system, through a data transformation application. The FAIRness of the output of the method, the FAIRified data and metadata, was evaluated using the FAIR Evaluation Services.

**Results:** We successfully applied our FAIRification method to the VASCA registry. Data entered on eCRFs is automatically transformed into machine-readable data and can be accessed and queried using SPARQL queries in the FAIR Data Point. Twenty-one FAIR Evaluator tests pass and one test regarding the metadata persistence policy fails, since this policy is not in place yet.

**Conclusion:** In this study, we developed a novel method for de-novo FAIRification via an EDC system. Its application in the VASCA registry and the automated FAIR evaluation show that the method can be used to make clinical research data FAIR when they are entered in an eCRF without any intervention from data management and data entry personnel. Due to the generic approach and developed tooling, we believe that our method can be used in other registries and clinical trials as well.

## 1. Introduction

The body of knowledge in medicine is growing exponentially [1] and the majority of publications in medicine are based on studies that put significant effort in the collection of data. However, it is estimated that 80% of the data that is collected in research cannot be reused, and is “reuseless” [2]. This is partially due to the fact that most datasets are not machine-actionable, nor machine-readable [2]. Data that are machine-actionable can be resolved by web services [3]), whereas machine-readable data are data in a format that can be automatically read and processed by a computer [4]. However, in our experience, the process of making data more machine-readable and machine-actionable is a major obstacle for data management personnel and researchers. This could lead to a focus on human-readability and findability. In this study, we show an approach that incorporates the process of making data machine-readable upon collection in an Electronic Data Capture (EDC) system.

### FAIR Data Principles

In 2016, a diverse group of researchers published the FAIR Data Principles [5]. These principles state that (research) data should be Findable, Accessible, Interoperable, and Reusable, for computers (machine-actionable and machine-readable) as well as for humans [5]. When data are made FAIR, it allows for more efficient use of the collected data, it improves the reproducibility of the data collection, and the data can more easily be reused for the same or other purposes than the initial data collection purpose. Possible applications of FAIR data include improved querying of research data [6] and interpreting and combining heterogeneous and challenging types of research data [7].

The FAIR Data Principles are getting increased traction. The original publication that proposed the principles is already cited over 2,500 times and the European Commission, as well as the National Institute of Health (NIH), are taking actions “to turn FAIR Data into reality” [8, 9]. Funders, such as the European Union’s (EU) Horizon 2020 [10] and The Dutch Research Council (NWO) [11], require researchers to put an effort into the FAIRification of their data (i.e., implementing the FAIR Data Principles in their research project) and document their FAIRification methodology into a Data Management Plan.

### FAIRification process

Jacobsen et al. proposed a general FAIRification workflow consisting of seven main steps [12]. First, one retrieves data that needs to be made FAIR and defines the FAIRification objective (i.e., why the data should be made FAIR and what data should be made FAIR). Next, one inspects the data and its current FAIR state (Figure 1.1) in order to determine challenges ahead for the next steps of the process. After that, the concepts and relations that describe the data are defined (Figure 1.2.1) and converted into a semantic data model: a canonical, machine-readable representation of data, based on ontology concepts (Figure 1.2.2). This applies to the data, as well as to the metadata that describe the registry as a whole and its access protocols. This resulting semantic data model is then used to transform the data into a machine-readable form. Considering that most ontologies have a representation in W3C-recommended Semantic Web technologies with the Resource Description Framework (RDF) as base data model, it is reasonable to use RDF to make the data linkable and ensure machine-readability (Figure 1.3). After that, one assigns a license for the usage of the data and assigns metadata (e.g., information about the research project and dataset) to it. Lastly, one makes their data and metadata available, e.g., by uploading the dataset with the corresponding metadata to a data repository or by serving a queryable access point on a web server. The dataset can then be found and queried, possibly as part of a federated query that spans multiple FAIR datasets.

**Figure 1:**
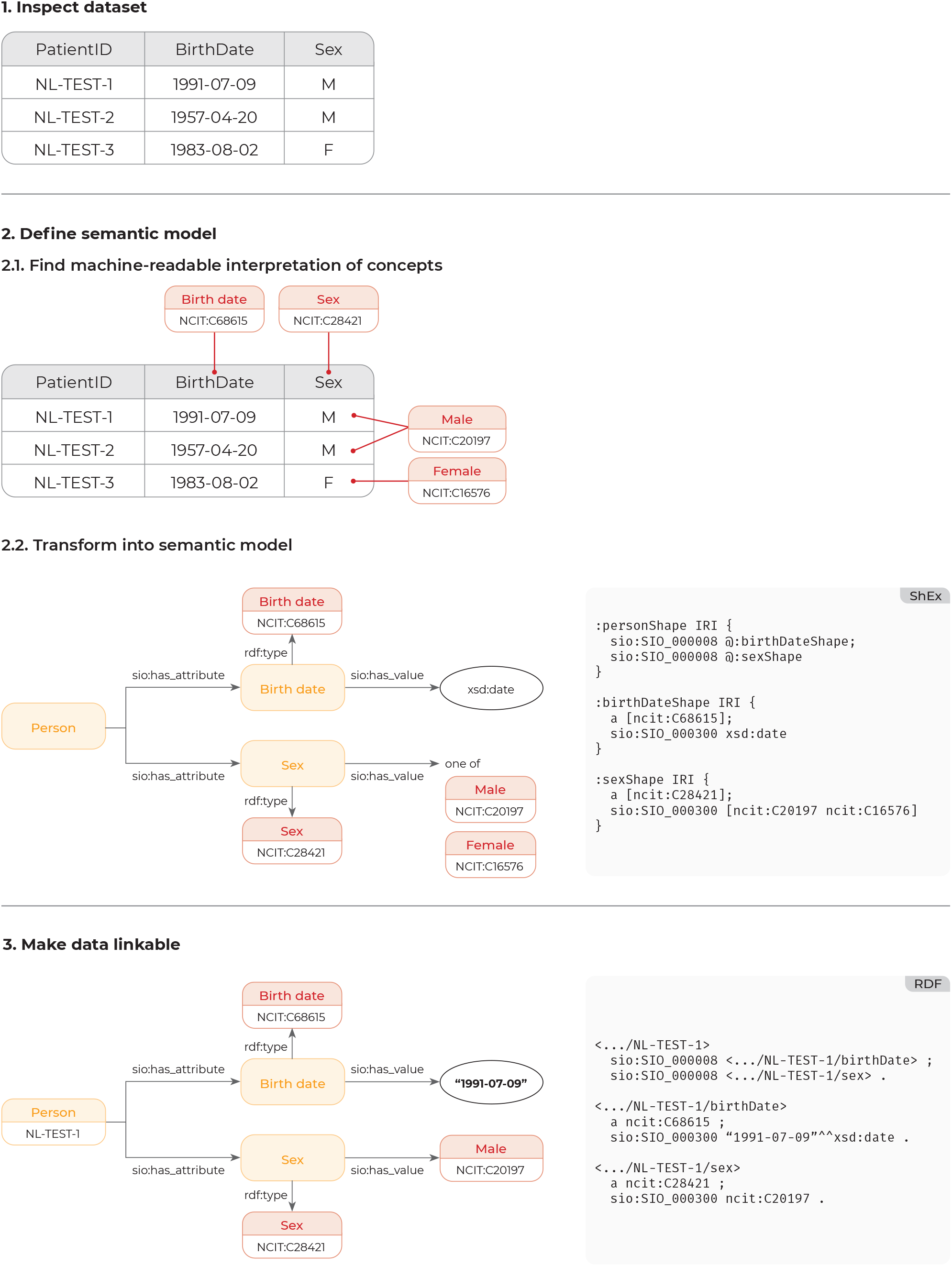
Schematic representation of the first three FAIRification steps.

### De-novo FAIRification via an EDC system

In this study, we introduce a novel method for de-novo FAIR-ification via an EDC system, the place where medical research data is often collected and stored via electronic Case Report Forms (eCRFs). Existing FAIRification workflows are usually carried out in a post-hoc manner: after the research project is conducted and data are collected in the EDC system (Figure 2.1). All the steps in this workflow have to be carried out by a FAIRification team in a semi-manual, labor-intensive manner. Post-hoc workflows can be partially reused for other research projects, but typically still require a FAIRification team. In [13] we described a workflow for de-novo FAIRification, where the FAIRification steps are incorporated in the process of setting up and collecting data for a registry or research project (Figure 2.2). Moreover, if a large part of the de-novo FAIRification workflow can take place in an EDC system, the reusability and, thus, scalability of FAIRification across research projects can be significantly improved.

**Figure 2:**
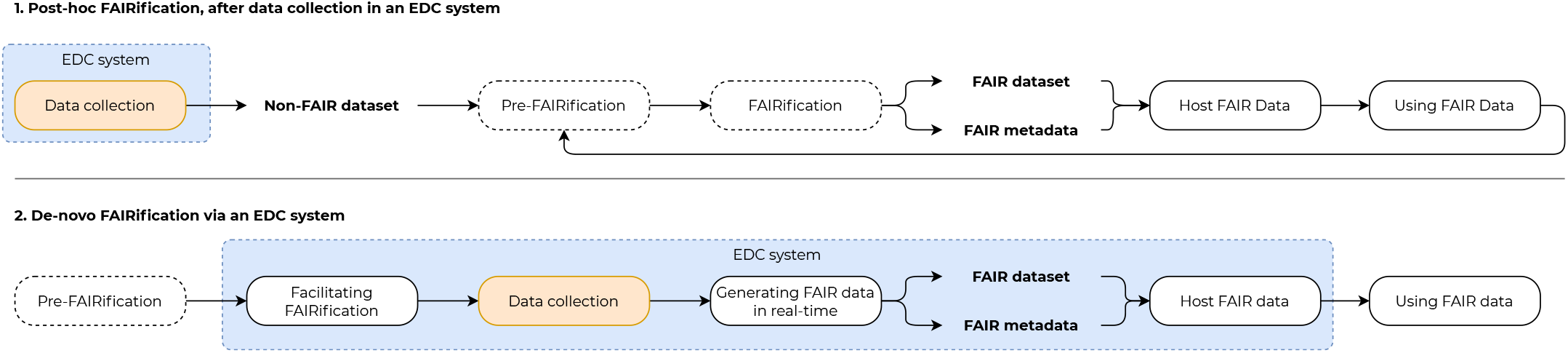
Post-hoc (1) and de-novo FAIRification (2) with an EDC system (adapted from [12] and [13], respectively). The EDC system takes care of most de-novo FAIRification steps.

To our knowledge, an application of FAIRification via an EDC system with a focus on machine-readable data has not been reported yet. Our method encapsulates four of the five de-novo FAIRification phases: facilitating FAIRification, data collection, generating FAIR data in real-time, and hosting FAIR data (highlighted in blue in Figure 2.2). After the FAIRification process and research project are set up (Facilitating FAIRification), data entry personnel can enter data in the EDC system (Data collection). The EDC system then takes care of generating (Generating FAIR data in real-time) and storing (Host FAIR data) machine-readable, FAIR data. For the last two steps, no intervention is needed from people involved in the medical research project. Moreover, FAIRification of data via an EDC system facilitates the querying of data directly from the sites where data are being collected, and thus avoids the creation of data silos and the need for data copying and centralization. To facilitate scalability in an environment where multiple EDC systems are used, it is important that FAIRification methods are not tied to a specific project or specific EDC system. In summary, to enable de-novo FAIRification at source by an EDC system, we identify five main requirements that are listed in Box 1.

#### Box 1

**Requirements for de-novo FAIRification via an EDC system**

1. The method should be EDC system and research project independent
2. The method should not require additional (technical) knowledge from end-users
3. Entered eCRF data can be transformed to a machine-readable format
4. The transformed data can be exposed and queried
5. The data and metadata are exposed in a manner following the FAIR Data Principles

### Case study: rare disease registry data

This study uses the Registry of Vascular Anomalies (VASCA), a rare disease (RD) registry that is part of the European Reference Network (ERN) on Rare Multisystemic Vascular Diseases (VASCERN), as a case study for de-novo FAIRification via an EDC system. As RDs by definition have a low prevalence [14], it is essential to combine and analyze data from various sources and create coherent, integrated datasets to have enough power to perform valid analyses. Making registry data FAIR would allow for virtual pooling of data across different registries, resulting in more data that can be analyzed in order to enhance the development of better diagnostic tests and therapies [15]. In an effort to increase the interoperability of RD registries, the EU’s Joint Research Center (JRC) produced a set of Common Data Elements (CDEs) to be adopted by RD registries [16]. In addition, VASCERN expressed the desire to build a virtual central registry by connecting multiple source registries that have implemented the FAIR Data Principles at source for that purpose. VASCA, therefore used the JRC CDEs to build eCRFs that will be used to collect data from Vascular Anomaly patients in nine countries, all in Electronic Data Capture platforms that follow the FAIR Data Principles.

In this study, we developed and implemented a method for de-novo FAIRification via an EDC system that automatically transforms eCRF data into machine-readable, FAIR data by the use of a semantic data model, a data transformation application, and a FAIR Data Point [17, 18]. We applied our method to the VASCA registry, in order to show its value in practice. The method is evaluated by assessing its impact on the Findability, Accessibility, Interoperability, and Reusability of the entered data using the FAIR Evaluation Services [19]. This article reports on the technical implementation and evaluation of the denovo FAIRification workflow, the entire workflow is described in detail in Groenen and Jacobsen et al [13].

## 2. Methods

Our method for de-novo FAIRification via an EDC system (Figure 3) consists of seven steps: 1) design eCRF, 2) implement semantic data model, 3) map eCRF structure to semantic data model, 4) transform data into RDF, 5) host FAIR data, 6) perform authentication and authorization, and 7) view, export, or query data. In this section, we describe these steps in detail. In order to support de-novo FAIRification via an EDC system, EDC vendor Castor EDC developed a data transformation application that transforms eCRF data into machine-readable data.

**Figure 3:**
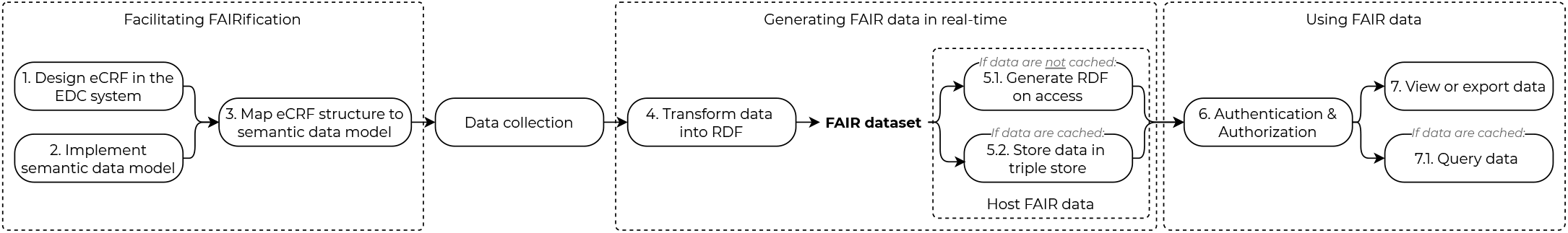
Simplified overview of the developed method for de-novo FAIRification via an EDC system.

### 2.1. Data transformation application

The data transformation application was developed using PHP 7.4 and Symfony 5.2 (backend), and React 16.13.1 (frontend). The data are stored in a MySQL 8 database. ARC2 2.5.1 was used as a triple store (i.e., a database that stores data in triples: subject - predicate - object). The data transformation application leverages the EDC system’s Application Programming Interface (API) [20] to get the eCRFs from the EDC system and uses the EDC system’s OAuth 2.0 implementation for authorization and authentication.

### 2.2. Development of method

#### 2.2.1. Design eCRF in the EDC system

The eCRFs, based on the CDEs provided by JRC, were implemented in a database in the EDC system Castor EDC [21]. The eCRF questions that we formulated according to the CDEs can be found in [22]. Some questions are repeated, such as the questions related to the patient’s genetic diagnosis: a patient can, for example, have one or more genetic diagnoses. The overview in [22] also lists if these questions are repeated, for the eCRF implementation of repeated questions (1:n relationship) differs from questions that are asked once (1:1 relationship) in the EDC system. More information about the eCRF implementation of the CDEs can be found in the publication that describes the setup of the VASCA registry [13].

#### 2.2.2. Implement semantic data model

The CDE elements and the relationships between them were captured in the semantic data model that can be accessed via [23]. We applied global ontologies and the W3C-recommended Resource Description Framework (RDF) to make our data and metadata machine-readable, considering that most biomedical ontologies are modeled in the RDF-based Web Ontology Language or are available in this form. RDF is built on web technologies that have proven to scale globally. We implemented the data model in the data transformation application, by adding all the different parts of the data model (groups, i.e., sets of triples - e.g. “Patient status”), elements (nodes, e.g. “Status”), and relationships between elements (triples, e.g. “Patient – has attribute – Status”) of the semantic data model. Figure 4 visualizes the difference between groups, nodes, and triples.

**Figure 4:**
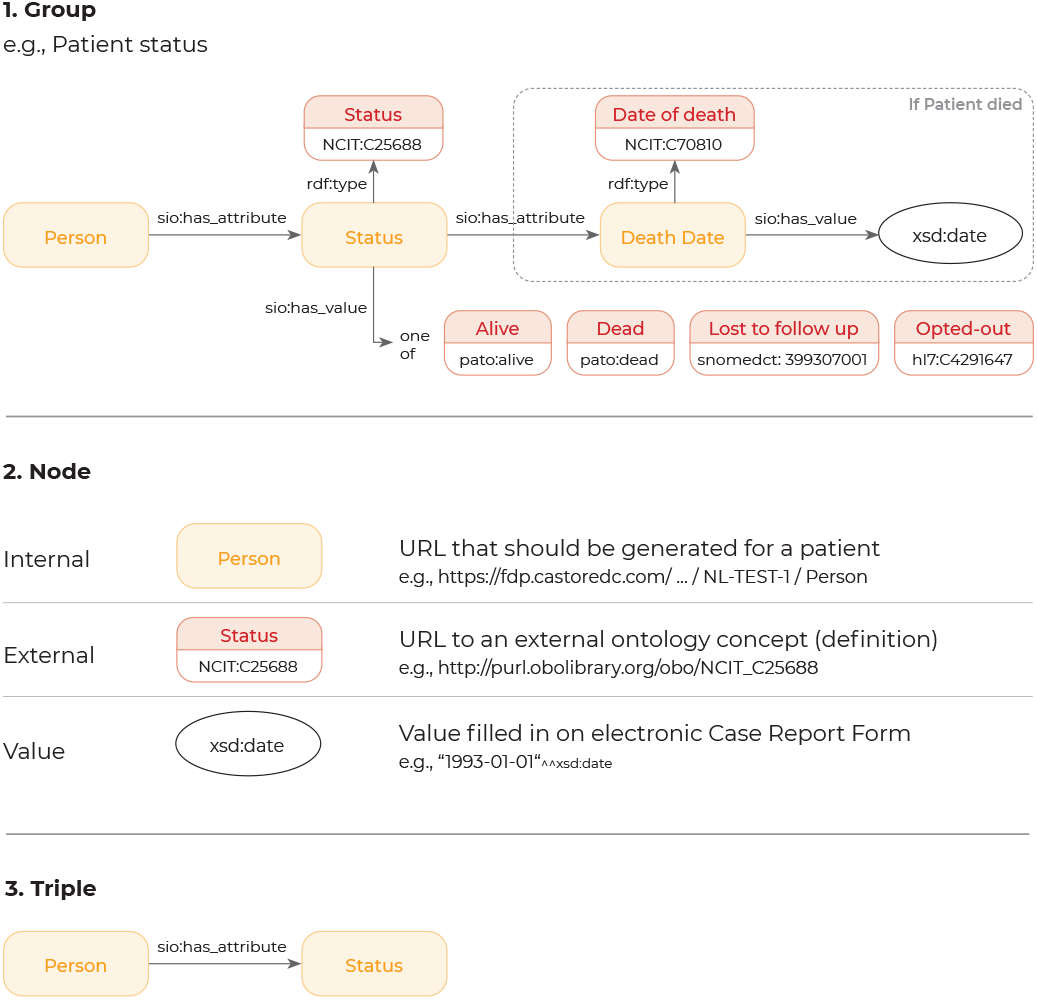
Examples of a group, node, and triple Groups (1) are smaller parts of the semantic data model and consist of a set of triples (3). Triples define the relationship between nodes (2) in the data model.

##### Groups

A group is a smaller part of the semantic data model and consists of a set of triples. The group can be marked as repeated (a 1:n relationship, e.g., a patient can have multiple genetic diagnoses) and dependent (e.g., the information about a patient’s diagnosis should only be included if a patient has a diagnosis).

##### Nodes

A node can be marked as an internal, external, or value node. An internal node is a URL that should be generated for a patient (e.g., https://fdp.castoredc.com/ … / [Patient Identifier] / [Node Name]). In Figure 4.2, Person is an internal node, Status an external node, and xsd:date (“1993-01-01”) a value node. Value nodes are linked to specific RDF data types and eCRF question types, as shown in Table 1. In the case of the example in Figure 4.1, a URL will be generated for every patient for the Person node and a value from the eCRF is expected in the value node.

**Table 1:** Relations between data transformation application node types, RDF data types, and eCRF question types

| Node type | RDF Data Type | eCRF question type |
| --- | --- | --- |
| Number | float, double, decimal, integer | Numeric, Radio button, Dropdown, Checkbox, Slider |
| Date and time | dateTime, date, time, gDay, gMonth, gYear, gYearMonth, gMonthDay | Date, Year, Time, Date and time |
| String | string | Radio button, Dropdown, Checkbox, Text, Text area |
| Boolean | boolean | Radio button, Dropdown, Checkbox |
| Annotated | URI | Radio button, Dropdown, Checkbox |

##### Triples

A triple is a combination of two nodes (subject and object) and a defining relationship between them (predicate). In Figure 4.3, the nodes Person and Status are related by the has attribute relationship: a Person has an attribute that is Status.

#### 2.2.3. Map eCRF structure to semantic data model

In order to transform the eCRF data according to the semantic data model, the model has to be mapped to the specific eCRFs and the questions on them.

##### Map model to variables

Nodes that have to be filled in with eCRF data (value nodes) have to be mapped to the specific questions (elements) on the eCRF. The mappings are made in the data transformation application. The application will only show eCRF questions that are compatible with the data type of the value node (e.g., a Year value node can only be mapped to an eCRF question of type Year or Date).

##### Map model to repeated forms

Some CDEs, and thus some eCRFs and parts of the semantic data model, can be repeated (a 1:n relationship in the eCRF, e.g., a patient can have multiple genetic diagnoses). In order to generate RDF for these repeated parts, the repeated form (eCRF) has to be mapped to the part of the semantic data model that has to be repeated.

#### 2.2.4. Data transformation

During the data transformation step, all records (patients) are retrieved from the EDC system using the system’s API. The transformation algorithm will then get the eCRF data of every record using the same API. The algorithm will then loop over all the groups in the data model. If the group is repeated, the algorithm will get every instance of the form that is mapped to the group (as described in step 3.2) and will render every group for those instances. If the group is dependent, the algorithm will determine if the group should be rendered based on the dependency clauses. If the group should be rendered, the algorithm will loop over all the triples in the group. For every triple, the URI of the subject node will be generated and the value will be retrieved from the EDC system if the object node is a value node. Every subject, predicate, and object will be added to a record-specific graph.

#### 2.2.5. Host FAIR data

The semantically modeled data can either be generated ‘on-the-fly’ when a user tries to access the data or cached by storing them in a triple store. During the development of the data transformation application, the generate-on-access approach was used to test the transformation of the data. After the development, the cache method was used. We expect that in most applications of our method, the data will be stored in a triple store to support querying.

##### Generate on access

For this storage approach, the RDF will be generated when a user tries to access the semantically modeled data. The data are stored in the EDC system only and the EDC system will handle authorization. It is not possible to query the data using SPARQL queries, because the data are not stored in a triple store, and retrieving the data is slower, since the data of all patients have to be loaded and transformed every time the RDF is accessed.

##### Cache

When the semantically modeled data are cached in a triple store, each record-specific graph (as described in step 4) will be stored in the triple store. For performance reasons, the application assesses for every record if it should be imported and thus if a graph has to be generated. If the record was created after the previous import, the record will be imported. If the record was updated after the previous import, the old graph will be archived and the record will be imported. If the record is not changed, it will not be imported. The data can be queried, because the triple store features a standard query interface for RDF (a ‘SPARQL endpoint’). Retrieving the data is fast, because the generation process already took place. Both the data transformation application and the EDC system handle authentication and authorization, because the data are present in both systems.

#### 2.2.6. Perform authentication and authorization

If the semantically modeled data are not publicly available, the user has to log in to get access to the data. Data will only be shown or returned to the user if they have access to the eCRFs of the specific institute (hospital) in the EDC system.

#### 2.2.7. View, export, or query data

The semantically modeled data can be accessed through a FAIR Data Point hosted by the EDC system. A FAIR Data Point is a metadata repository that provides access to metadata of digital objects in a FAIR way [17]. The metadata are modeled by the Data Catalogue Vocabulary version 2 (DCAT2), a W3C-recommended semantic model for describing catalogs [24]. Conform DCAT2, the data are available as a ‘distribution’ (i.e., a specific representation of a dataset [24]) in the FAIR Data Point and users can view or export the data by clicking the buttons in the user interface, or by programmatically navigating through the URLs that are listed in a machine-readable description of the distribution. When the data are cached in the triple store, they can also be queried using SPARQL queries. Metadata for the catalogs, datasets, and distributions are publicly available. Based on the data access policy, users have to authenticate themselves before they can access the semantically modeled data (step 6).

### 2.3. Evaluation of method

We evaluated the output of the method, the FAIRified data and metadata, using the Evaluator from the FAIR Evaluation Services [19]. The Evaluator registers, assembles, and applies community-relevant sets of Compliance Tests against a digital resource, in this case the FAIRified data and metadata, and provides a detailed report about what a machine “sees” when it visits that resource [19]. We ran the “All Maturity Indicator Tests as of May 8, 2019” set of tests against the endpoints from the Catalog, Dataset, and Distribution layers of the FAIR Data Point. The result of every test contains the outcome (status) of the test (passed test: “success”, failed test: “failure”) and a log of the execution of the test.

## 3. Results

Our method for de-novo FAIRification via an EDC system was applied to the VASCA registry. Here we describe the application of the method to the registry and the results of the (automated) evaluation.

### 3.1. Application of method

We describe our results based on the steps of our developed FAIRification method (Figure 3).

#### 3.1.1. Design eCRF in the EDC system

Seven CDEs (Pseudonym, Personal information, Patient status, Care pathway, Disease history, Diagnosis, Research) were implemented on an eCRF in Castor EDC. The eCRF questions related to these CDEs are listed in [22]. Figure 5 shows a filled eCRF in the EDC system. The CDE for Disability was not added for logistic reasons, as described in [13].

**Figure 5:**
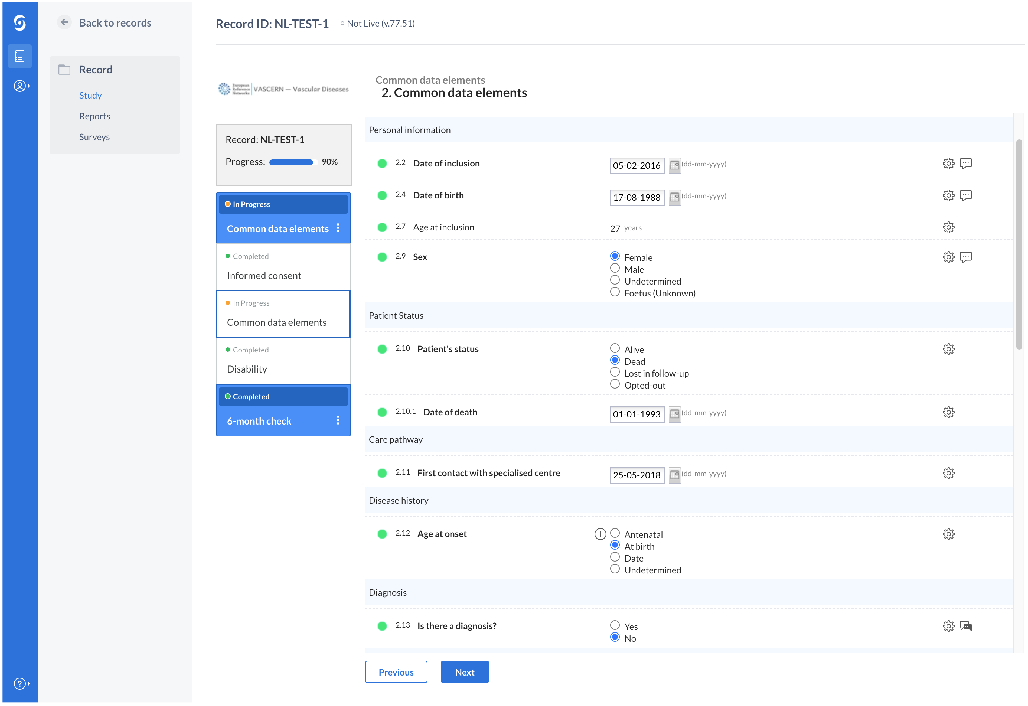
Screenshot of an eCRF in the EDC system.

#### 3.1.2. Implement semantic data model

The semantic data model with its eleven modules (groups) [23] was added to the data transformation application. The Genetic Diagnosis and Undiagnosed (phenotype of patient) modules from the model were entered as repeated groups in the application. Figure 6.1 shows the triples related to the Patient Status group and Figure 6.2 shows a visual representation of the group’s nodes and the relationships between them. The semantic data model was versioned in the application (version 0.1.0, [23]), such that new versions of and mappings to the model can be added when needed.

**Figure 6:**
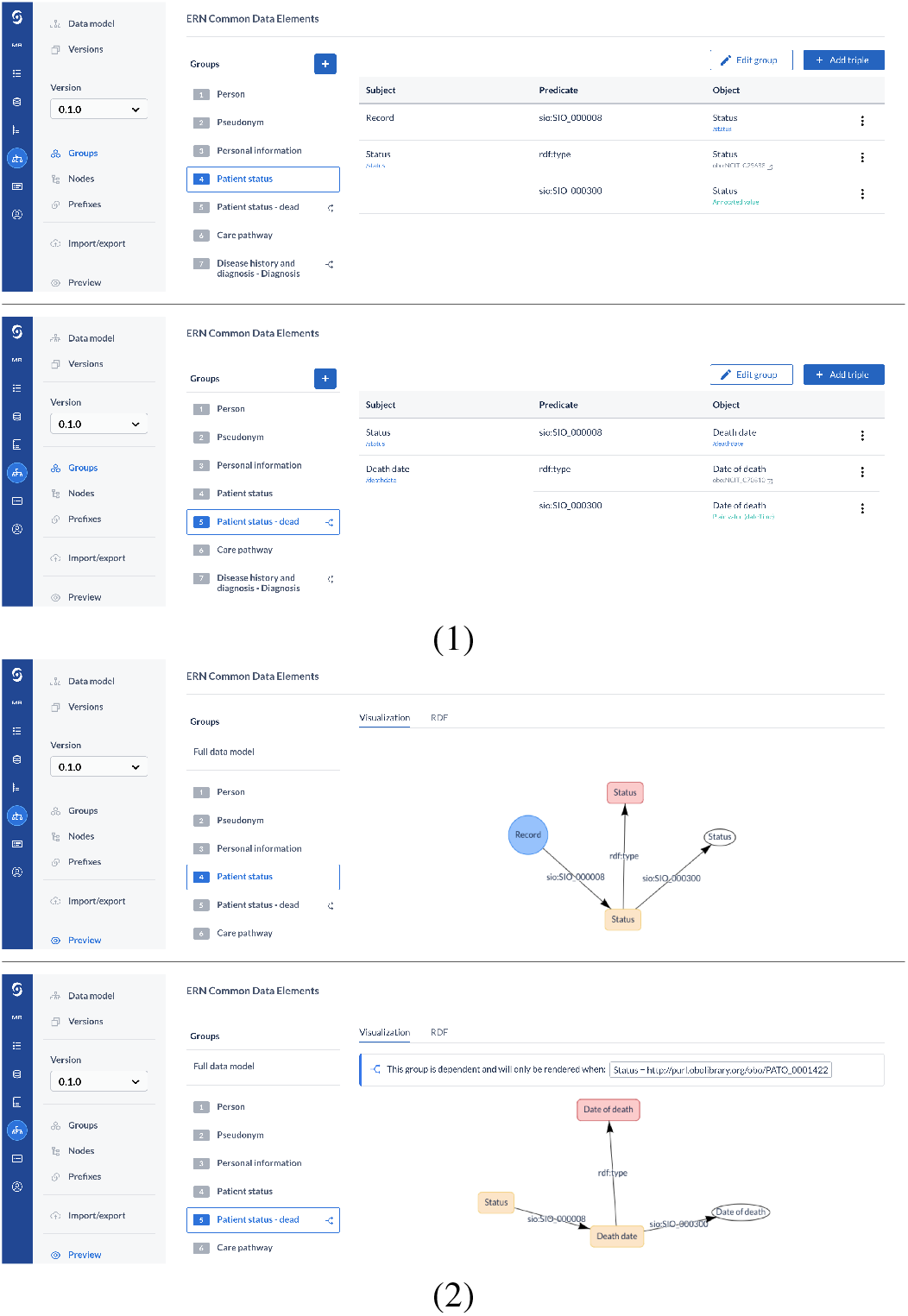
Triples related to the Patient Status group (1) and a visual representation of these triples (2) in the data transformation application.

#### 3.1.3. Map eCRF structure to semantic data model

The mappings between the questions on the eCRF and the semantic data model were added in the data transformation application. Figure 7.1 shows an overview of all mappings. Figure 7.2 shows the process of mapping the Status value node to the ‘Patient’s status’ question on the eCRF and Figure 7.3 shows the process of mapping the values of this question to ontology concepts.

**Figure 7:**
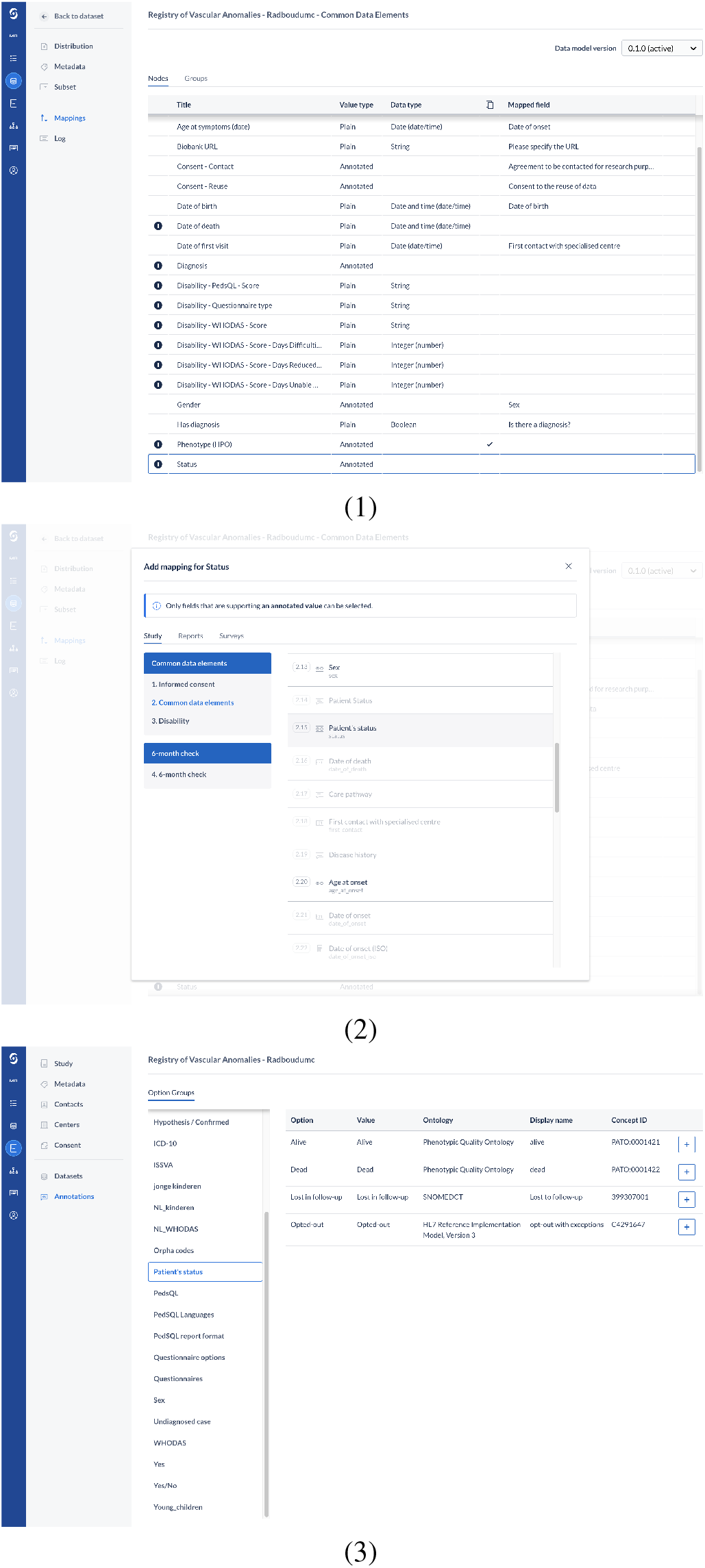
The overview of all mappings (1), mapping of an annotated value node to an eCRF variable (2), and annotation of the eCRF values with ontology concepts (3) in the data transformation application.

#### 3.1.4. Transform data into RDF

The transformation of the data is currently performed periodically. With a predefined interval, every six hours, the data are pulled from the EDC system and changes are updated in the RDF. A method to support the transformation of the data based on a trigger (e.g., entering data on the eCRF) is being explored. Figure 8 shows a part of the generated RDF related to the status of the patient and the meaning of the individual lines in the RDF.

**Figure 8:**
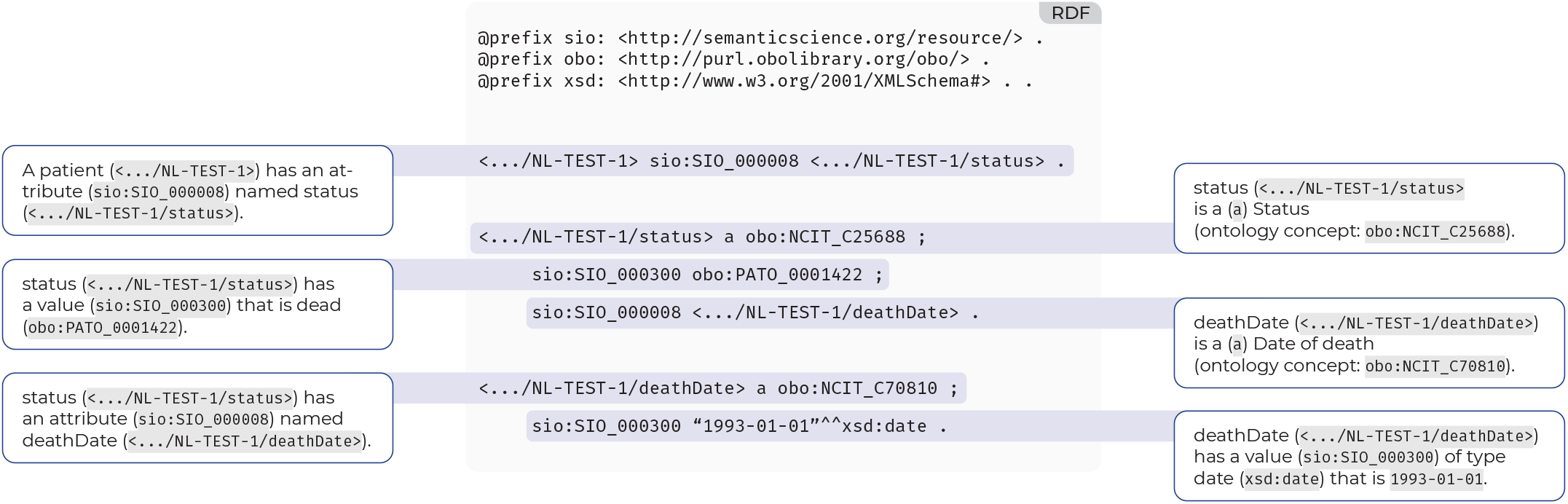
Generated RDF related to the Patient Status group.

#### 3.1.5. Host FAIR data

We piloted the data transformation using the previously described generate-on-access approach. After we determined that the RDF that was generated using the data transformation application was correct and thus following the semantic data model specification, we switched to the cached approach. As described above, we generate and cache the data on an interval.

#### 3.1.6. Perform authentication and authorization

When users try to access the data via the FAIR Data Point, they are redirected to the EDC system to authenticate themselves. The EDC system determines if the user has access, based on access policies defined by institutions in the system, and redirects the user back to the FAIR Data Point. Authentication and authorization currently work by using the user interface of the FAIR Data Point.

#### 3.1.7. View, export, or query data

The data and metadata are accessible through the FAIR Data Point of the EDC system [25]. The FAIR Data Point provides a user interface for browsing the (meta)data visually, and also a machine-readable endpoint to perform this task automatically. The Registry of Vascular Anomalies is available as a separate catalog (i.e., a collection of metadata about resources [24], in this case: registry datasets). Every center contributing to the registry has its own dataset in this catalog and, therefore, also defines its own data access policy. In the user interface, the user is given the option to view and export the RDF or query the data of a specific distribution per dataset (i.e., a specific representation of the dataset [24], in this case: an RDF representation of the CDE data). An example query is shown in Figure 9. Querying of data across different distributions (federated querying) can be performed through external SPARQL clients. We are currently working on a query interface in the FAIR Data Point that supports federated querying as well.

**Figure 9:**
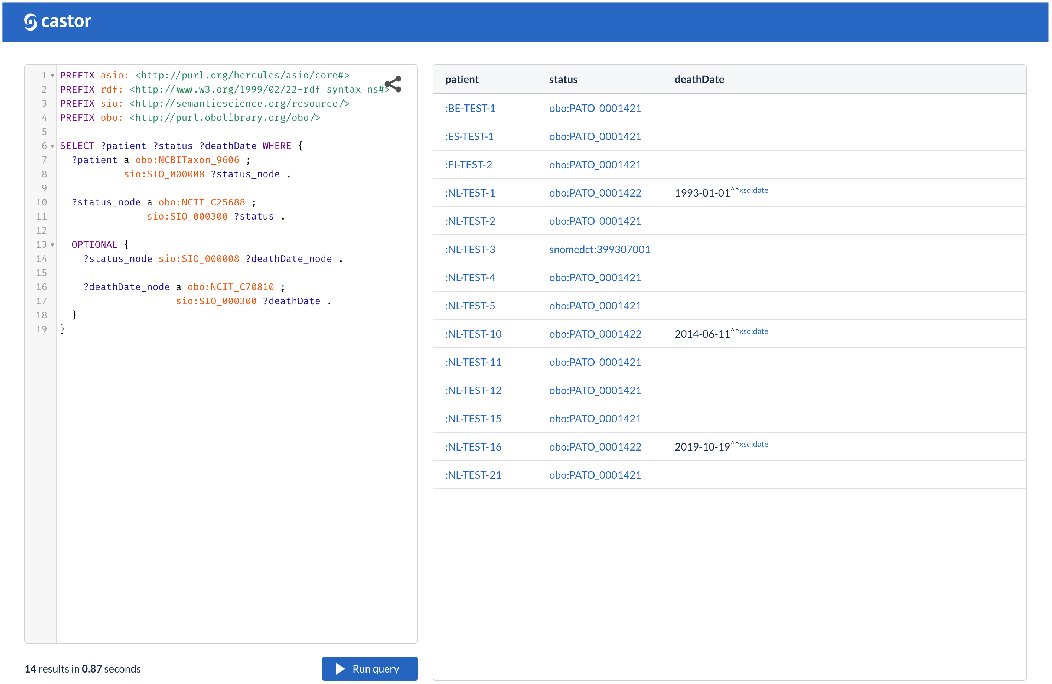
SPARQL query in the FAIR Data Point listing the identifier (patient), status (status), and a possible date of death (deathDate) of patients.

### 3.2. Assessment of the output of the method

We assessed the FAIRness of the output, the transformed data, by using the FAIR Evaluator. We found that initially, per FAIR Data Point layer, fourteen tests passed and eight tests failed (Table 2). Five out of eight tests regarding Findability passed, two out of five tests regarding Accessibility, five out of seven tests regarding Interoperability, and two out of two tests regarding Reusability. The FAIR Data Principles associated with the tests can be found in Box 2.

**Table 2:**
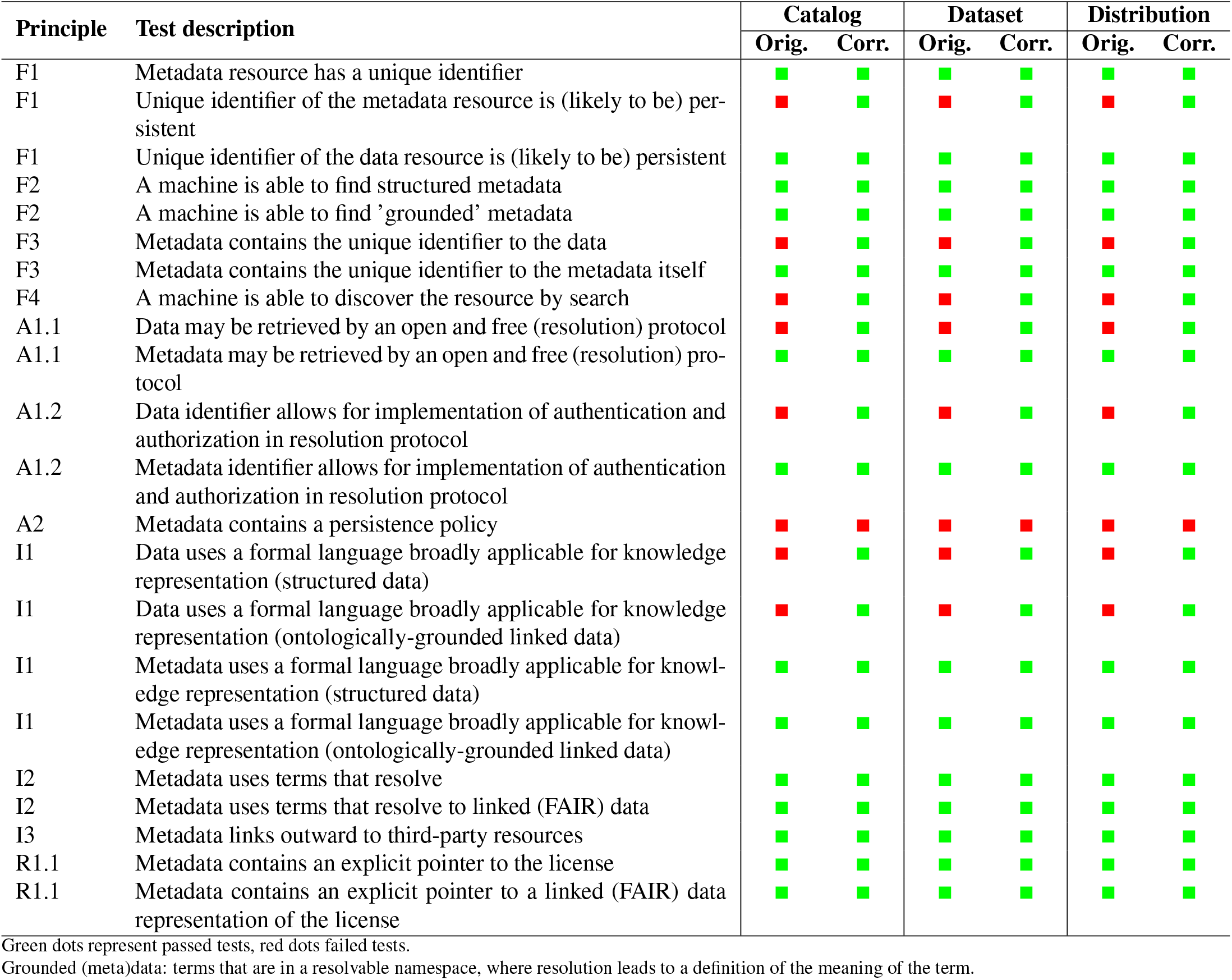
Original (Orig.) and Corrected (Corr.) outcomes of the FAIR Evaluator

There were three main reasons why the eight tests failed (Table 3): 1) the FAIR Evaluator was unable to find the associated data (F1, F3, A1.1, A1.2, I1), 2) the metadata could not be found in Bing (F4), and 3) there was no persistence policy in place (A2). The tests associated with the first and second reasons should have passed, based on our interpretation of the description of either the test or the FAIR Data Principle associated with the test. The first reason is a known limitation of the Evaluator: the Evaluator does find the predicate that links to the distribution (dcat:distribution), but does not follow links inside of RDF documents and therefore cannot find the predicate that links to the data (dcat:accessURL). The metadata and data that are exposed in the FAIR Data Point can indeed not be found in Bing (reason 2), but can be found in Google and the FAIR Data Point Registry [26]. Therefore a machine is still able to discover the resources in our FAIR Data Point by a search. We, therefore, corrected the outcome of seven tests (as marked in Table 3). With these corrections, 21 tests pass and one test fails (Table 2). The failing test (A2) indicates that we should define a persistence policy to make sure that the metadata can still be accessed when the data are not available anymore.

**Table 3:**
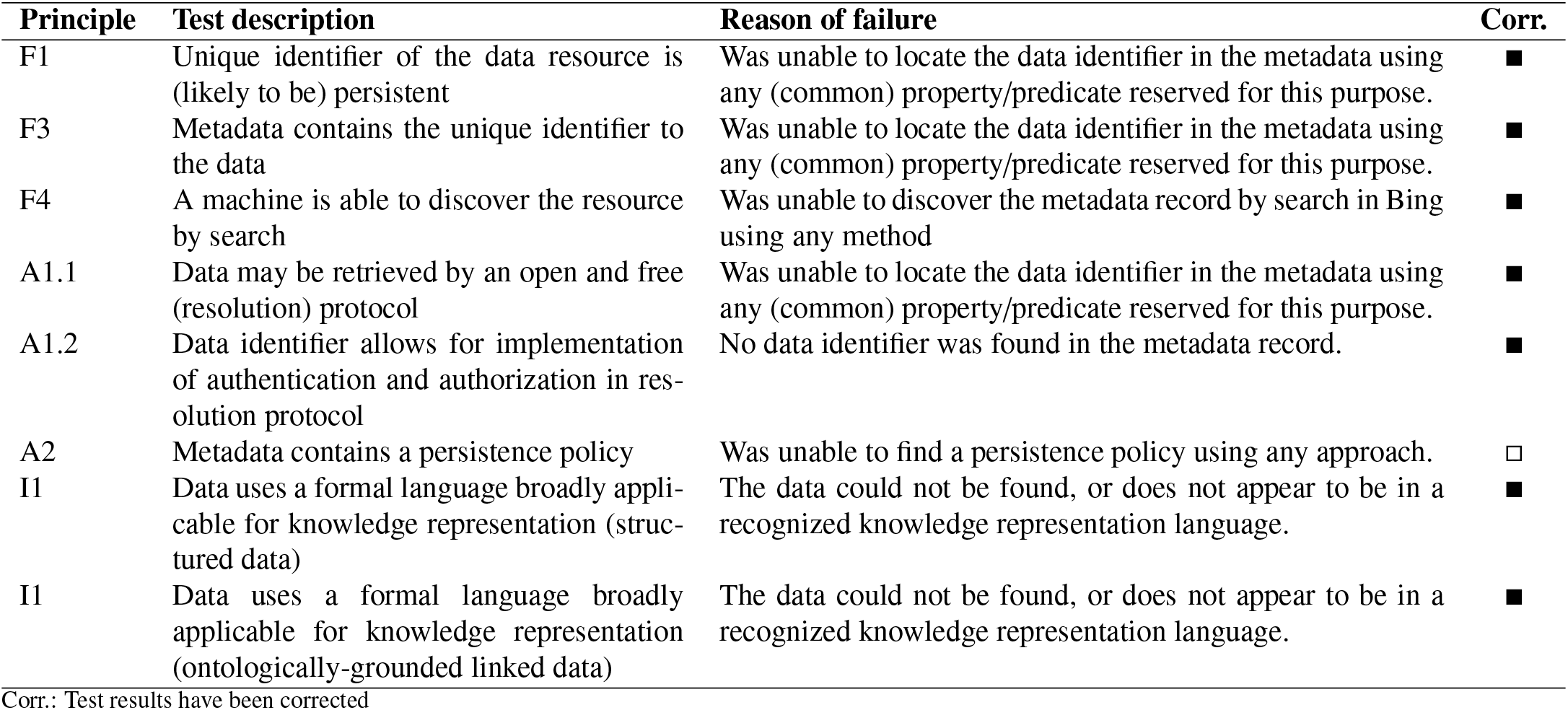
Failed tests and the reason of their failure

| Principle | Test description | Reason of failure | Corr. |
| --- | --- | --- | --- |
| F1 | Unique identifier of the data resource is (likely to be) persistent | Was unable to locate the data identifier in the metadata using any (common) property/predicate reserved for this purpose. | ■ |
| F3 | Metadata contains the unique identifier to the data | Was unable to locate the data identifier in the metadata using any (common) property/predicate reserved for this purpose. | ■ |
| F4 | A machine is able to discover the resource by search | Was unable to discover the metadata record by search in Bing using any method | ■ |
| A1.1 | Data may be retrieved by an open and free (resolution) protocol | Was unable to locate the data identifier in the metadata using any (common) property/predicate reserved for this purpose. | ■ |
| A1.2 | Data identifier allows for implementation of authentication and authorization in resolution protocol | No data identifier was found in the metadata record. | ■ |
| A2 | Metadata contains a persistence policy | Was unable to find a persistence policy using any approach. | □ |
| I1 | Data uses a formal language broadly applicable for knowledge representation (structured data) | The data could not be found, or does not appear to be in a recognized knowledge representation language. | ■ |
| I1 | Data uses a formal language broadly applicable for knowledge representation (ontologically-grounded linked data) | The data could not be found, or does not appear to be in a recognized knowledge representation language. | ■ |

Corr.: Test results have been corrected

## 4. Discussion

In this study, we developed and applied a method for de-novo FAIRification via an EDC system. Our method particularly focused on the steps to make eCRF data actionable and readable by machines. We defined five main requirements (Box 1). Below we describe the insights that we gained in addressing these requirements.

The data model mapping step in our method makes sure that the method is EDC and research project independent (requirement 1). Data models are not hard coded in the data transformation application, but can be added by a FAIR data expert in a user-friendly interface and then can be mapped to questions on the eCRF by the person that is building the eCRF. This does not require any technical knowledge (requirement 2). The data transformation application is a separate application outside of the EDC system and therefore other EDC systems with an API can be integrated as well. Our method ensures that data that is entered into the eCRF is automatically made FAIRer, because the data and metadata are automatically transformed into a machine-readable format and exposed in a FAIR Data Point (requirements 3 and 4). After the data model is entered in the data transformation application and mapped to the eCRF, the application does not need any intervention from data entry or data management personnel. The developed method is used in the VASCA registry to automatically generate machine-readable data from data entered on the registry’s eCRFs.

### Box 2

**FAIR Data Principles, as described in [5]**

**F1** (Meta)data are assigned a globally unique and persistent identifier

**F2** Data are described with rich metadata (defined by R1 below)

**F3** Metadata clearly and explicitly include the identifier of the data they describe

**F4** (Meta)data are registered or indexed in a searchable resource

**A1** (Meta)data are retrievable by their identifier using a standardized communications protocol

**A1.1** The protocol is open, free, and universally implementable

**A1.2** The protocol allows for an authentication and authorization procedure, where necessary

**A2** Metadata are accessible, even when the data are no longer available

**I1** (Meta)data use a formal, accessible, shared, and broadly applicable language for knowledge representation.

**I2** (Meta)data use vocabularies that follow FAIR principles

**I3** (Meta)data include qualified references to other (meta)data

**R1** (Meta)data are richly described with a plurality of accurate and relevant attributes

**R1.1** (Meta)data are released with a clear and accessible data usage license

**R1.2** (Meta)data are associated with detailed provenance

**R1.3** (Meta)data meet domain-relevant community standards

### 4.1. Impact on the data’s FAIRness

One of the requirements of our method was to meet as many FAIR principles as possible (requirement 5). Below we revisit the four FAIR facets, Findability, Accessibility, Interoperability, and Reusability, and their associated principles (Box 2), and discuss our method’s impact on them.

#### 4.1.1. Findability

As shown in Table 2, all tests regarding the Findability of the data and metadata pass. By using an implementation of the FAIR Data Point, we ensure that the data are assigned a persistent identifier (F1). The distribution is assigned a unique identifier and linked to the metadata (F1, F3). Every patient in the EDC system also receives a unique identifier (F1). The FAIR Data Point provides a metadata scheme (F2) and method for storing metadata and the data they relate to [17] (F4).

#### 4.1.2. Accessibility

All FAIR Evaluator tests, despite the persistence policy check, pass (Table 2). The metadata in the FAIR Data Point can be accessed through a REST API and the data through exporting the RDF or querying the data using SPARQL. REST and SPARQL are both standardized communication protocols (A1, A1.1). An authentication and authorization layer is present: the end-user has to log in to the EDC system before the data are returned to them (A1.2). We are exploring options for implementing an authentication and authorization method for when the data are accessed by machines. A policy for providing access to the metadata, even when the data are not available anymore, has to be defined in the future (A2). Implementing the latter will also make sure that the only corrected FAIR Evaluator test that currently failed, will pass.

#### 4.1.3. Interoperability

The tests regarding the interoperability of the data pass as well (Table 2). By transforming the eCRF data to machine-readable data (in RDF) that follows an established semantic data model with links to ontology concepts, we comply with the first and second Interoperability principles (I1, I2). The use of the FAIR Data Point also ensures that we use appropriate vocabularies for the description of metadata and the data they relate to (I2).

#### 4.1.4. Reusability

As described above, we use the FAIR Data Point’s metadata schema, with attributes from the Data Catalog Vocabulary (DCAT2) [24] and Dublin Core (DC) Terms [27] (R1, R1.3) and a semantic data model with links to community-accepted ontologies (R1.3). Both of the tests regarding the licensing of the (meta)data (Table 2), therefore, pass. The provenance for the data and metadata can be specified in the FAIR Data Point (R1.2), with a person responsible for the (meta)data and a person that can be contacted for inquiries about the (meta)data attached to every dataset and distribution. For the VASCA registry, there is no data usage license available yet (R1.1). We are exploring methods to allow for requesting access to the data. The support for a data usage license will be a part of this and will be added to the FAIR Data Point, once developed.

### 4.2. Strengths and limitations

The main strength of this study is the EDC and research project independent approach we took. Since the data transformation application and its integration with the EDC tool (Castor EDC) are generic, the application can potentially also be used for other registries or clinical trials in the same EDC or in other EDCs. The method for de-novo FAIRification via an EDC system only requires a one-time setup: once the semantic data model and mappings to the eCRF questions are entered into the application, the application will automatically make the data machine-readable when entered into the EDC system. This eases a burden of FAIR data experts. They only have to be actively involved in the FAIRification when the research project is being set up, with the exception of data model changes, instead of being actively involved with FAIRification throughout the entire project. Moreover, there is no change in workflow from the point of view of data management or data entry personnel. They do not have to consider the FAIRification workflow during the development of eCRFs and data collection, while others can immediately reuse the machine-readable data.

The start of our FAIRification method is still quite technical. Entering the semantic data model into the data transformation tool and adding the mappings between the model and the eCRF requires some technical expertise. We aim to simplify this process in the future by allowing for the import of the model or by automating the mapping process. The semantic data model used in this study was already created and we, therefore, did not describe the steps needed to create such a model. However, creating a semantic data model is a complex task. We recommend that users of our FAIRification method gather expertise from domain and semantic data modeling experts, which is the first step of the de-novo FAIRification workflow [13].

### 4.3. Comparison to other literature

This paper, to our knowledge, is one of the first that incorporates the entire FAIRification workflow in the process of a medical research project and makes data FAIR via an EDC system, which is often used as a source for data collection in medical research. There are, however, other studies that describe partial implementations of the FAIR workflow in medical research.

Sinaci et al. published a FAIRification workflow for health research [28] based on the post-hoc workflow from GO FAIR [29]. Their workflow takes a raw dataset as starting point, rather than our approach where the data is collected and the dataset is created during the FAIRification workflow. The Open Source Registry System for Rare Diseases (OSSE), also software that is used for electronic data capture, has included parts of the FAIRification workflow in their software, such as exposing metadata in a FAIR Data Point [30].

Other related work has focused on making data more interoperable. Wolstencroft et al. created an application that allows scientists to annotate data present in Excel spreadsheets with ontology concepts [31], which can be incorporated in the post-hoc workflow. Pang et al. developed a system for semi-automatically matching entered data to ontology concepts, upon data collection [32]. Lastly, the work of Sernadela et al. [33] is also closely related to our work. Sernadela et al. have created a method to harmonize data across patient registries using a semantic web layer to allow for federated querying. Their approach is a post-hoc approach, where the data are made FAIR registry by registry, after they are collected. Our method also allows for data harmonization across registries and research projects, however, in our case, the data is made FAIR upon collection, which significantly improves the method’s scalability.

### 4.4. Design decisions

Several design decisions were made during the project. The three most important decisions were decisions regarding the generation of RDF, storage of the data, and authentication and authorization.

#### 4.4.1. Generation of RDF

To make sure that the initiators of a research project and data management personnel can be more closely involved in the FAIRification process, we aimed for a method for the generation of RDF that did require limited to no prior knowledge of linked data. In the pilot phase of the project, we implemented an RDF generation method that used Twig (Box 3), a templating language [34] where parts of an RDF document were placeholders for the data that could be wrapped in conditional blocks (e.g., only show the date of death triples if the patient has died). This method proved that data could be extracted from the eCRF and could be transformed into RDF, but also had some disadvantages. While implementing the data model, we noticed that the Twig template became harder to read and maintain, since almost every item of the data model was wrapped in a conditional block. In addition, creating and maintaining Twig templates still requires substantial knowledge of programming languages, which most initiators and data management personnel limitedly have.

##### Box 3

**Example Twig syntax for the status of the patient**

~~~
 <{{ record.URL }}/> sio:SIO_000008
        <{{ record.URL }}/status>.
 <{{ record.URL }}/status> a obo:NCIT_C25688 ;
        sio:SIO_000300 {{record.status}}.
 {% if record.status == “obo:PATO_0001422” %}
        <{{ record.URL }}/status> sio:SIO_000008
               <{{ record.URL }}/deathDate>.
        …
 {% endif %}
~~~

Placeholders (in brackets {{ … }}) will be replaced with data from the eCRF.

Therefore, we switched to the model-based approach as described in the Methods section. The semantic data model can be built once in the data transformation application by a FAIR data expert and, subsequently, data management personnel can map their eCRF structure to this model. In this way, data management personnel do not have to write code themselves and making their eCRFs linkable, by mapping their structure, only takes them a few minutes.

#### 4.4.2. Storage of the data

For storing the data, we identified two main database options: a graph database (that supports triple stores and querying out of the box) and a SQL or relational database (that integrates with our PHP framework out of the box). We decided to use a MySQL database for the storage of the data model and mappings. A MySQL-based triple store was used for the storage of the transformed eCRF data. MySQL databases were chosen since the EDC system’s infrastructure was built around them and because we had limited knowledge of using triple stores for storing and altering data. This also makes sure that the EDC system can provide support for the data transformation application and it allows for possible integration of the application in the EDC system.

#### 4.4.3. Authentication and authorization

To ensure that only authorized users can access the transformed data, we had to implement an authentication and authorization layer in the FAIR Data Point. We decided to use the EDC system’s authentication and authorization mechanism for now, since that allowed us to ensure that the access conditions that are currently set in the EDC system are followed. We are planning to look into other Authentication and Authorization Infrastructures (AAIs) to also allow non-EDC users to get access to the data. The latter will enable other registries and ERNs to query the data using a federated approach.

### 4.5. Implications for practice

Machine-readable and machine-actionable data allows for the combination of data of various sources, thus allowing for the analysis of more data. In the case of rare disease registry data, this is highly beneficial, since data in every single source is limited due to the low prevalence of rare diseases. Using our approach, data are automatically made FAIR when entered on the eCRF, without putting a burden on the data management and data entry personnel involved in the registry. The data transformation application can be reused for other (RD) registries and clinical trials, due to the generic mapping approach.

### 4.6. Future work

We are still in the process of improving the user interface of the data transformation application in order to make it available for other clinical trials and registries. In addition, we are looking into automated mapping algorithms to assist the end-user with mapping the value nodes to the eCRF questions. A method for federated querying should also be implemented to support querying across different distributions, and in the case of the VASCA registry, across different centers. Last, we are exploring methods for machine-readable data use conditions and requesting access to the data when a user does not have access to the registry data in the EDC. This also includes integrating with AAIs as an alternative to the authentication and authorization with the EDC system.

## 5. Conclusion

We successfully developed a method for de-novo FAIRification via an EDC system that automatically transforms eCRF data entered into the system into machine-readable, FAIR data by the use of a semantic data model and a data transformation application. Our method is currently used in the VASCA registry.

Its application shows that de-novo FAIRification via an EDC system can be used to make clinical research data FAIR when they are entered on an eCRF without any intervention from data management or data entry personnel. The developed method can be applied to and reused in other (RD) registries and clinical trials, due to the generic approach and developed tooling.

## Data Availability

Not applicable

## Abbreviations

AAI: Authentication and Authorization Infrastructures
API: Application Programming Interface
CDE: Common Data Element
DC: Dublin Core
DCAT2: Data Catalogue Vocabulary version 2
eCRF: electronic Case Report Form
EDC: Electronic Data Capture
ERN: European Reference Network
EU: European Union
FAIR: Findable, Accessible, Interoperable, and Reusable
JRC: Joint Research Center
NIH: National Institute of Health
NWO: The Dutch Research Council
RD: rare disease
RDF: Resource Description Framework
VASCA: Vascular Anomalies
VASCERN: European Reference Network on rare vascular diseases

## CRediT authorship contribution statement

**Martijn G. Kersloot:** Conceptualization, Methodology, Software, Writing - Original Draft. **Annika Jacobsen:** Conceptualization, Methodology, Writing - Review & Editing. **Karlijn H.J. Groenen:** Conceptualization, Methodology, Writing Review & Editing. **Bruna dos Santos Vieira:** Writing - Review & Editing. **Rajaram Kaliyaperumal:** Writing - Review & Editing. **Ameen Abu-Hanna:** Supervision, Writing - Review & Editing. **Ronald Cornet:** Conceptualization, Supervision, Writing - Review & Editing. **Peter A.C. ‘t Hoen:** Writing - Review & Editing. **Marco Roos:** Conceptualization, Supervision, Writing - Review & Editing. **Leo Schultze Kool:** Conceptualization, Supervision, Writing - Review & Editing. **Derk L. Arts:** Conceptualization, Supervision, Writing - Review & Editing.

